# Clinical course of COVID-19 patients admitted to the Intensive Care Unit of a tertiary care hospital in Central India

**DOI:** 10.1101/2023.06.07.23290823

**Authors:** Alka Modi Asati, Rakesh Patel, Kritika Singhal, Chakresh Jain

**Affiliations:** Department of Community and Family medicine, AIIMS, Bhopal; Department of Medicine, SSMC Rewa

**Author notes:** **Corresponding author** Dr. Alka Modi Asati., Address: Department of Community and Family Medicine, 2^nd^ floor, Sardar Vallabhai Patel Bhawan, AIIMS, Saket Nagar, Bhopal, MP-462020. **Author approval** All the authors have seen and approved the manuscript. **Competing interests** There are no competing interests. **Funding statement** This study had no source of funding.

**Keywords:** COVID -19, Covid mortality, comorbidity, ICU Outcome, Pneumonia, Tertiary care

## Abstract

**Background:** COVID-19 pandemic also known as Corona virus pandemic, is caused by severe acute respiratory syndrome coronavirus 2 (SARS-CoV 2). In India, the first case of COVID-19 was reported on 30 January 2020 while in Madhya Pradesh on 20 March 2020 and in Rewa on 27 April 2020. Nearly 95 % of people recovered from COVID-19, and nearly three to five percent of cases needed Intensive Care Unit care and most of them needed mechanical ventilation.

**Materials and Methods:** This was a hospital based cross-sectional study, done among 75 clinical or RT-PCR confirmed cases of COVID-19 infection admitted to the ICU of tertiary care unit.

**Results:** In the present study, 63% were male and maximum (35%) belonged to 41-60 years of age. The most common symptom was fever at the time of admission to the hospital. Co-morbidity was reported in 21(28%) of patients. Out of these, majority of patients recorded combination of hypertension and diabetes as the most common comorbidity.

**Conclusion:** Delayed medical intervention, advanced age, and the presence of underlying health conditions, such as cardiovascular disease, diabetes, etc., are known risk factors for severe illness and can contribute to worse outcomes and increased mortality in COVID-19 patients.

## Introduction

The COVID-19 pandemic has affected millions of people globally, with many requiring hospitalization and intensive care unit (ICU) treatment. ICU treatment is necessary for patients with severe COVID-19 symptoms, including respiratory failure, pneumonia, and other complications. This infection started in China in December 2019 and became a pandemic, caused by severe acute respiratory syndrome coronavirus 2(SARS-CoV 2). (1) In India, the first case of COVID-19 was reported on 30 January 2020.(2) first wave of the COVID pandemic in India lasted from January to November 2020., While a few cases kept appearing, the second wave evolved from February - July 2021. (3)

Till May 2021, more than 164 million confirmed cases and more than 3.4 million deaths attributed to COVID-19. (3) Nearly 95 % of people recovered from Covid-19, and around 3 to 5 percent of cases required ICU care (2) including mechanical ventilation. COVID-19 pandemic has affected many aspects of people’s life including physical, social, emotional and behavioral wellbeing. To halt the spread of SARS-CoV-2 infection and alleviate its health effects, country had enforced different control measures, like social distancing, partial and total lockdowns, closing schools and businesses and wearing face masks. Although such measures have helped in flattening the epidemic curve, a COVID-19 resurgence has been reported after resuming all public activities. This increased morbidity and mortality associated with COVID-19 especially during the second wave.(2,4) The COVID-19 vaccine development was an important step towards controlling the pandemic. India’s COVID Vaccination drive was started on January 16, 2021, in a phased manner. (5) Even after all this, many people were seriously affected and were admitted in hospitals, especially during the second wave. Most of these patients were admitted to the Intensive Care Units and would eventually require invasive mechanical ventilation (MV) because of diffuse lung injury and acute respiratory distress syndrome (ARDS). Most of the ICU reports from the United States have shown that severe COVID-19-associated ARDS (CARDS) is associated with prolonged MV and increased mortality.(6) There may be institutional and regional variations in ICU outcomes and mortality. The unsatisfactory ICU outcomes and high mortality observed during CARDS have raised worries about the use of mechanical ventilation. India, now with the largest population in the world, had difficulty in treating severe COVID-19 cases because the country had only 49,000 ventilators (6). This disproportionate availability of resources led to different types of outcomes in COVID-19 patients, this study throws light on the clinical course of COVID-19 patients in the tertiary care units with the objective of clinical characteristics and outcomes of COVID-19 patients admitted to ICU.

## Material & Methods

This study was a hospital based cross sectional study which was conducted for a period of one month in May 2021. Universal sampling technique was applied to select the participants. All the patients admitted to the ICU of a tertiary care hospital were approached and consent was taken from patients or their attendant. A total of 75 RT-PCR positive or clinically diagnosed (CT scan findings suggestive of COVID) patients were included in the final analysis. The study tool was a structured proforma, which was designed, validated and used for data collection. Information was collected from various sources including treatment sheet, patients, and their attendant. All patients admitted to ICU for COVID-19 treatment, and more than 12 years of age were included in the study. Data related to socio-demography, clinical characteristics and laboratory parameters were collected.

### Statistical analysis

Continuous data were presented as median and IQR, Frequency and percentages (n; %) were used for categorical variables. Appropriate test applied wherever necessary. Analysis was done with R software (4.3.1) and Microsoft excel.

### Ethical consideration

The study was conducted after taking ethical approval from the institutional ethical committee of Sham Shah Medical College, Rewa, Madhya Pradesh. This study was approved by the committee with permission letter numbered IEC/MC/2020470. Confidentiality was maintained by removing identifiers during analysis and reporting of results. Data was collected only after due informed consent/assent.

## Result

Out of 75 patients admitted to the ICU, mean age of the patients was 53.66 (±17.19) years. Maximum number of participants (35%) belonged to the 41-60 years age group followed by 21-40 years (31%). Most of the study participants were residents of rural area. Sociodemographic characteristics are presented in Table 1.

**Table 1:** Socio-demographic characteristics of patients.

| **Sociodemographic Characteristic** | **N = 75 (%)** |
| --- | --- |
| **Age** |  |
| <20 | 6 (8.0%) |
| 20-40 | 23 (31%) |
| 41-60 | 26 (35%) |
| >60 | 20 (27%) |
| **Sex** |  |
| Female | 28 (37%) |
| Male | 47 (63%) |
| **Locality** |  |
| Rural | 49 (65%) |
| Urban | 26 (35%) |

Out of 75, only 48 (64%) patients gave the history of close contact with any COVID-19 positive individual and 68% of the patients were found positive for COVID-19 infection. Although 32% patients were COVID-19 negative, but their clinical status was strongly suggestive of COVID-19 on CT scan. Table 2 depicts the clinical characteristics of the participants.

**Table 2:**
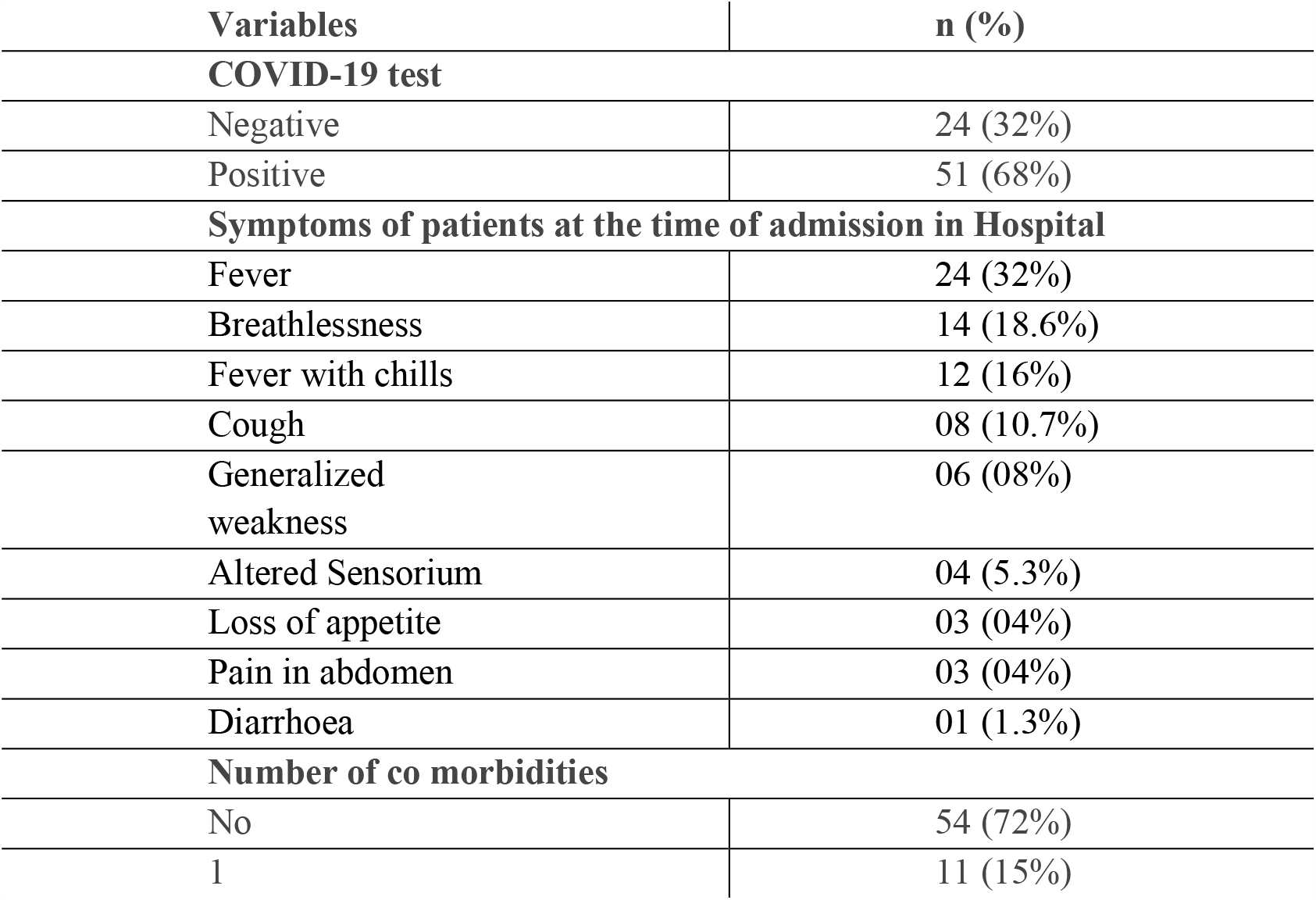
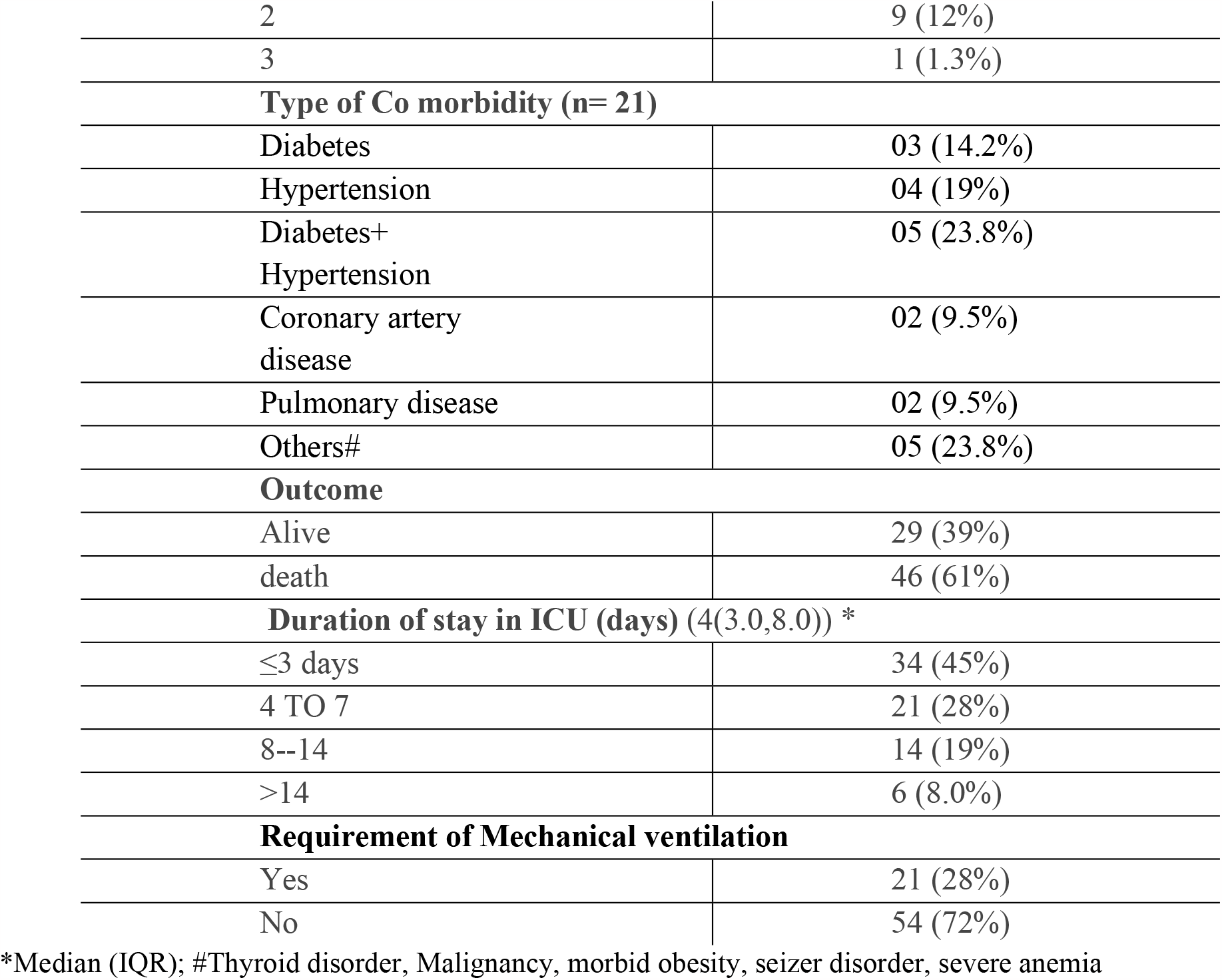
Clinical profile and outcome of patients admitted in ICU.

| **Variables** | **n (%)** |
| --- | --- |
| **COVID-19 test** |  |
| Negative | 24 (32%) |
| Positive | 51 (68%) |
| **Symptoms of patients at the time of admission in Hospital** |  |
| Fever | 24 (32%) |
| Breathlessness | 14 (18.6%) |
| Fever with chills | 12 (16%) |
| Cough | 08 (10.7%) |
| Generalized weakness | 06 (08%) |
| Altered Sensorium | 04 (5.3%) |
| Loss of appetite | 03 (04%) |
| Pain in abdomen | 03 (04%) |
| Diarrhoea | 01 (1.3%) |
| **Number of co morbidities** |  |
| No | 54 (72%) |
| 1 | 11 (15%) |
| 2 | 9 (12%) |
| 3 | 1 (1.3%) |
| **Type of Co morbidity (n= 21)** |  |
| Diabetes | 03 (14.2%) |
| Hypertension | 04 (19%) |
| Diabetes+<br>Hypertension | 05 (23.8%) |
| Coronary artery<br>disease | 02 (9.5%) |
| Pulmonary disease | 02 (9.5%) |
| Others# | 05 (23.8%) |
| **Outcome** |  |
| Alive | 29 (39%) |
| death | 46 (61%) |
| **Duration of stay in ICU (days) (4(3.0,8.0)) \*** |  |
| ≤3 days | 34 (45%) |
| 4 TO 7 | 21 (28%) |
| 8--14 | 14 (19%) |
| >14 | 6 (8.0%) |
| **Requirement of Mechanical ventilation** |  |
| Yes | 21 (28%) |
| No | 54 (72%) |

\*Median (IQR); #Thyroid disorder, Malignancy, morbid obesity, seizure disorder, severe anemia

Symptoms during admission to the hospital, were fever (32%), followed by breathlessness (18.6%) and cough (10.7%). In the current study, around 28% of patients had any comorbidity. Out of that 52.4% of patients experienced at least one comorbidity (Table 3). Hypertension (19% patients reported) was most common comorbidity followed by diabetes (14.2%) whereas hypertension and diabetes were the most common (23.8%) combination comorbidity reported. In view of duration of stay in ICU, most of the patients (45%) were admitted for less than 3 days to ICU. A total of 28% required mechanical ventilation. With respect to outcomes, 39% alive and 61% patients expired at the end of the study period.

**Table 3:** Laboratory investigations of patients at the time of admission in ICU:

| Parameter | N (%) |
| --- | --- |
| **Haemoglobin\*** | 12.00 (10.00, 13.65) |
| **Anaemia** |  |
| No | 30 (40%) |
| Mild | 18 (24%) |
| Moderate | 24 (32%) |
| Severe | 3 (4.0%) |
| **WBC\*** | 12,000 (9,300, 15,800) |
| **PLATELET\*** | 126,000 (112,000, 179,500) |
| **Platelet** |  |
| Normal | 59 (79%) |
| Thrombocytopenia | 16 (21%) |
| **CRP** |  |
| Negative | 28 (37%) |
| Positive (raised) | 47 (63%) |
| **D DIMER** |  |
| ≤0.5 | 8 (11%) |
| >0.5 | 67 (89%) |
| **HRCT SCORE\*** | 17.0 (14.5.0, 21.0) |
| <8 (Mild) | 01 (1.3%) |
| 8-15 (Moderate) | 28 (37.34%) |
| >15 (Severe) | 46 (61.34%) |
| **Disease severity status#** |  |
| Mild to moderate | 36 (48%) |
| Severe | 39 (52%) |

\* Median (IQR); #Disease severity status=Severe disease was defined as either of these, i.e., RR >24/min, SpO2 <94 per cent on room air, confusion, drowsiness, hypotension, sepsis, septic shock or admission to ICUs (7).

Table 4 shows association of outcome of patients with their sociodemographic and clinical characteristics. Maximum patients from age group > 60 years expired but this result was not significant. Only duration of stay in ICU and HRCT Score showed significant association with outcome. Figure 1 pictorially depicts the distribution of clinical and demographic characteristics across the clinical outcome through a stacked bar chart.

**Table 4:**
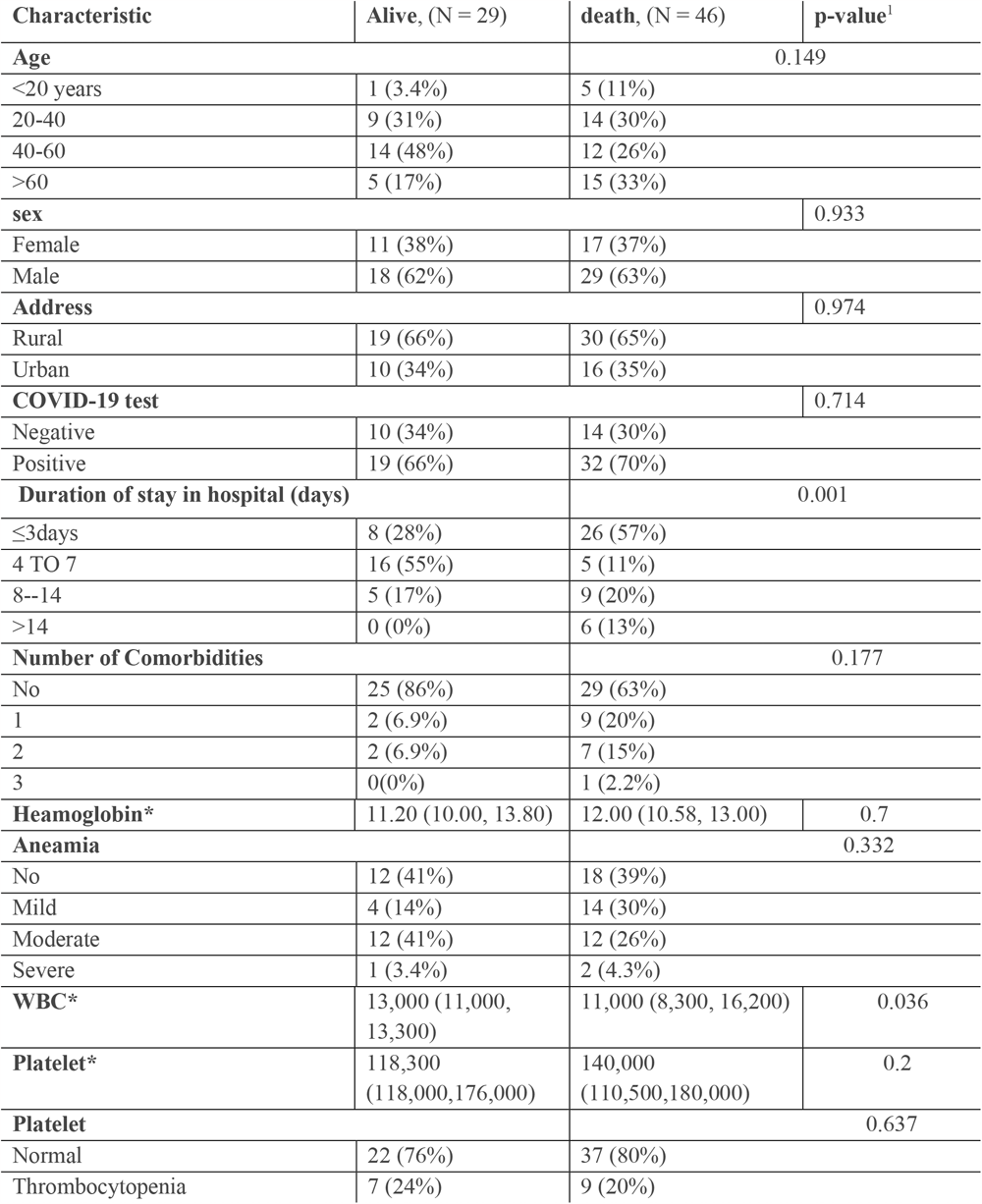
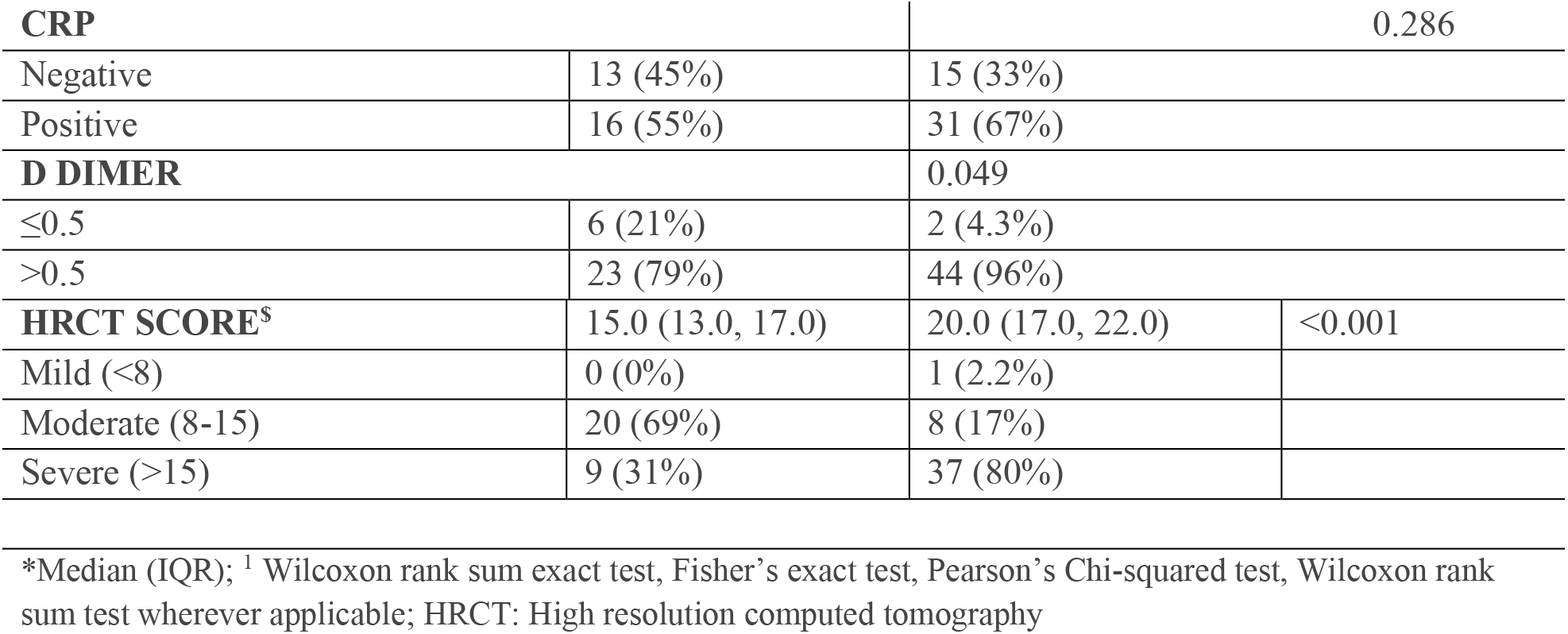
Distribution of patient outcomes across sociodemographic and clinical characteristics.

**Figure 1:**
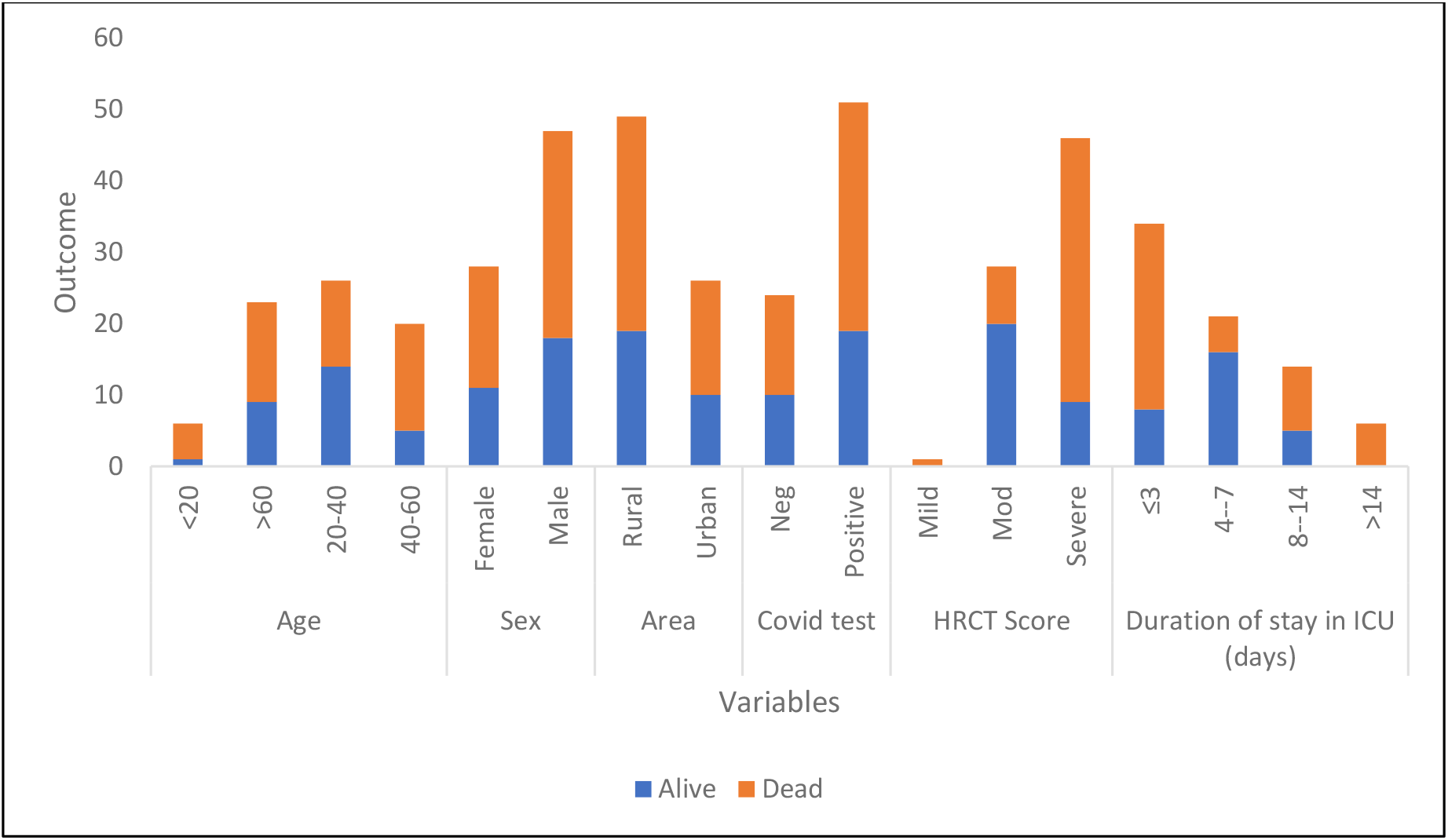
Clinical Outcome across sociodemographic and clinical characteristics.

## Discussion

Our study reported the sociodemographic characteristics, clinical profile, and outcomes of the 75 patients admitted to ICU for COVID-19 infection during the month of May 2021 in the tertiary care hospital. Compared with other studies, the mean age of study participants was almost similar i.e., 53.66 years in our study and 59 and 64 years in the study of Sweden and Atlanta respectively. (8,9) This study reported that age and gender were not significantly associated with higher mortality. Similar results were reported in a study conducted in Saudi Arabia. (10) However, a study conducted in Sweden reported that age was significantly and gender was not significantly associated with mortality. (7) Most of our study participants were male. Around 68.8% participants experienced at least one comorbidity in a study conducted by A. Assiri et al. (10) where as in our study only 28% patients experienced any comorbidity. Hypertension was most common comorbidity followed by diabetes. Similarly in a study conducted in Sweden hypertension (39.6%) and diabetes (26.2%) were found to be the most prevalent comorbidity. (8) Auld et al reported that hypertension (61.7%) was most common followed by diabetes (45.6%) .(9) The most prevalent symptom found was fever (32%), followed by breathlessness (18.6%) and cough (10.7%) at the time of admission to the hospital in the current study. Similar results were found in a study conducted in Nanjing, China where most common symptoms were fever, dry cough followed by dyspnea. Further, cough (34.7%) was the most common symptom followed by fever (17.4%) in a study conducted in North India. (7) In our study 28% required mechanical ventilation, in contrast to 76% and 83.2% respectively in a study conducted by Auld et al (8) and Oliveira E et al. (11) Inadequate resources, technical capacity and early deaths might be the reason for this low proportion of patients requiring mechanical ventilation. A study from Sweden reported that 30.3% patients died and 31.3% were discharged. (8) Similarly, a study from Atlanta reported that 28.6% patients died in the ICU, 67.7% were transferred out of the ICU and 3.7% remained in the ICU. (9) A Central Florida study reported that out of 131 patients admitted to the ICU, 80.2% remained alive at the end of the study period, 70.9 % were discharged from the hospital and 19.8% of patients expired. (11) In contrast, our study reported higher deaths in the ICU due to COVID-19, i.e.,61% died. Our study reported a higher mortality than other literature. The reason behind this high mortality may be a delay in arrival of patients to the health facility, delays in treatment due to overcrowded health facilities, exhausted healthcare workers, inadequate training, etc. Further, presentation of patients with severe infection or end stage disease might also be a reason. Around the world, ICU mortality due to COVID-19 ranged from 20–62%. (11) In view of the duration of stay in the ICU, maximum (48%) were admitted for less than or equal to 3 days. The mean duration of stay in ICU was less in our study than the studies conducted in Florida and Sweden. (7,10) This may be due to the facility being a tertiary care institute, so patients with severe symptoms or end stage disease were admitted in a higher proportion.

### Limitation

We conducted a study on only for one month and during the second wave of COVID-19 epidemic in India, hence consent was difficult to obtain due to high number of deaths. The result cannot be generalized as the study was conducted in a hospital setting.

### Recommendation

It is needed to empower the health facility to accommodate a cohort of critically ill patients in one location. Appropriate training related to triage of patients, use of personal protective equipment and development of Standard of procedures for treatment are required for providing proper care to patients.

## Conclusion

Second wave of COVID-19 -, presented with 41-60 years of age, male gender, and comorbidities as risk factor for severe disease and admission to ICU. Preventive measures should be emphasized on population with such risk factors to prevent COVID-19 and associated mortality in them.

## Data Availability

All data produced in the present study are available upon reasonable request to the authors.

## Notes

### Competing Interest Statement

The authors have declared no competing interest.

### Funding Statement

This study did not receive any funding

### Author Declarations

The study was conducted after taking ethical approval from the institutional ethical committee of Sham Shah Medical College, Rewa, Madhya Pradesh. This study was approved by the committee with permission letter numbered IEC/MC/2020470.

